# Functional brain network and trail making test changes after major surgery and delirium

**DOI:** 10.1101/2021.12.10.21267535

**Authors:** Simone JT van Montfort, Fienke L Ditzel, Ilse MJ Kant, Ellen Aarts, Lisette M Vernooij, Claudia D Spies, Jeroen Hendrikse, Arjen JC Slooter, Edwin van Dellen

## Abstract

**Background:** Delirium is a frequent complication of elective surgery in elderly patients, associated with an increased risk of long-term cognitive impairment and dementia. Disturbances in the functional brain network were previously reported during delirium. We hypothesized persisting alterations in functional brain networks three months after elective surgery in patients with postoperative delirium, and hypothesized that postoperative brain connectivity changes (irrespective of delirium) are related to cognitive decline.

**Methods:** Elderly patients (N=554) undergoing elective surgery underwent clinical assessments (including Trail Making Test B (TMT-B) and resting-state functional magnetic resonance imaging (rs-fMRI) before and three months after surgery. Delirium was assessed on the first seven postoperative days. After strict motion correction, rs-fMRI connectivity strength and network characteristics were calculated in 246 patients (130 patients underwent scans at both timepoints), of whom 38 (16%) developed postoperative delirium.

**Results:** Rs-fMRI functional connectivity strength increased after surgery in the total study population (β=0.006, 95%CI=0.000–0.012, p=0.021), but decreased after postoperative delirium (β=-0.014, 95%CI=0.000–0.012, p=0.026). No difference in TMT-B scores was found at follow-up between patients with and without postoperative delirium. Patients who decreased in functional connectivity strength declined in TMT-B scores compared to the group that did not (β=11.04, 95%CI=0.85-21.2, p=0.034).

**Conclusions:** Delirium was associated with decreased functional connectivity strength three months after the syndrome was clinically resolved, which implies that delirium has lasting impact on brain networks. Decreased connectivity strength was associated with statistically significant (but not necessarily clinically relevant) cognitive deterioration after major surgery, which was not specifically related to delirium.

**Summary statement:** Delirium was associated with decreased resting-state fMRI functional connectivity strength three months after the syndrome was clinically resolved. Irrespective of delirium, decreased connectivity strength after major surgery was associated with a statistically significant cognitive deterioration.

## Introduction

Delirium, a clinical expression of acute encephalopathy, is characterized by an acute disturbance in attention and awareness, with additional cognitive deficits.^1^ Delirium is by definition a consequence of one or more medical conditions and affects 15-25% of elderly patients after major elective surgery,^2^ in that case called postoperative delirium. Delirium can be accompanied with patients’ distress and increased length of admission, and is related to poor outcomes, such as long-term cognitive impairment and dementia.^2^

Functional connectivity (the statistical interdependencies of time-series recorded from different brain areas), for example measured with functional magnetic resonance imaging (fMRI), is thought to reflect functional interactions between brain areas that are crucial for cognitive functioning.^3^ The macroscale functional brain network, an abstraction of all communication pathways between remote brain regions, may be studied in relation to cognitive processing or pathological conditions.^4^ It has been hypothesized that delirium is a disconnection syndrome, reflecting a breakdown of functional brain networks.^5,6^

The overall functional connectivity strength is decreased in a range of neuropsychiatric disorders, including delirium.^6–13^ Moreover, the functional brain network organization in delirium is topologically altered, and characterized by a less efficient and less integrated organization.^10,11^ On a regional level, alterations in regional functional connectivity between parts of the default mode network (DMN, i.e. the posterior cingulate cortex, PCC) and the central executive network (CEN, i.e. the dorsolateral prefrontal cortex, DLPFC), were additionally shown in delirium patients compared to non-delirious controls.^14^ These specific interactions are of interest for delirium, because awareness and attention disturbances are core symptoms. The DMN is implicated in wakeful rest and non-task states, and its activity is negatively correlated with the CEN, which is involved in selective attention processing.^15^ Seven days after resolution of delirium, a decreased global functional connectivity strength has been observed, suggesting long-term impact of the disorder on the functional brain network.^10^ A decrease of global functional connectivity strength and disturbances in functional network efficiency and integration have also been reported in cognitive impairment and dementia.^13,16,17^

A decrease of global functional connectivity strength due to delirium may relate to impaired outcomes, such as long-term cognitive impairment or dementia. However, up to now, studies evaluating delirium in relation to the brain network lacked baseline and follow-up measurements, i.e. fMRI measurements before the development of delirium, and after its resolution.^10,14^ It is unknown whether delirium is related to a lasting change in global functional connectivity strength or brain network organization. The aim of this study was to test the hypothesis that postoperative delirium leads to a decrease in global functional connectivity strength, efficiency and integration of the functional brain network three months postoperatively compared to preoperatively.^10,11^ As a secondary analysis, the long-term effect of delirium on the regional connection between the PCC and the DLPFC was studied.^14^

## Materials and methods

### Study design and population

This study is part of the *Biomarker Development for Postoperative Cognitive Impairment in the Elderly* (BioCog) project at the University Medical Center (UMC) Utrecht and Charité Hospital at Berlin (ethical approval numbers 14-469 respectively EA2/092/14).^18^ Inclusion criteria were European ancestry, age of 65 years or above, scheduled for a major elective surgery (i.e. orthopedic-, cardiac-, gastro-intestinal-, gynecological, urologic, maxillofacial- or otorhinolaryngologic surgery) of at least 60 minutes, an fMRI measurement at baseline and/or follow-up, and signed informed consent. Patients with one or more of the following characteristics were excluded: a life expectancy shorter than a year or evidence for (early) dementia as indicated with a score of 23 or lower on the Mini Mental State Examination (MMSE).^19^ This study adheres to the STROBE (Strengthening the Reporting of Observational Studies in Epidemiology) guidelines.^20^

### Procedures

Included patients were invited to the hospital for a baseline measurement, i.e. a clinical assessment and an MRI scan of the brain. The clinical assessment was administered by trained research staff. After surgery the patient group was followed in the hospital twice daily for delirium assessments until the day of discharge, with a maximum of seven postoperative days. Approximately three months after surgery, the patients were invited to the hospital again for the three-month follow-up visit with similar measurements as during the baseline visit, i.e. a clinical assessment and an MRI scan of the brain.

### Clinical assessment

#### Trail making test

At baseline and at follow-up, the trail making test (TMT) was administered. We specifically focused on the TMT in this study, because decreased TMT test scores have previously been associated with delirium severity and also with decreased functional connectivity strength (measured with EEG) in dementia with Lewy bodies, a condition that is related to delirium.^16,21^ Visual memory is required for TMT section A and executive functioning are required for TMT section B.^22^

#### Other characteristics

Preoperative alcohol use was assessed using the self-reported Alcohol Use Disorders Identification Test (AUDIT).^23^ A cut-off value of 8 points was used to define alcohol misuse.^24^ Preoperative depressive symptoms were estimated with the self-reported Geriatric Depression Scale (GDS) with 15 items.^25^ A score of 6 was used as cut-off to define depression. Preoperative functional impairment was measured with the Barthel Index.^26^ Preoperative physical status was scored by anesthesiologists (in training), using the validated American Society of Anesthesiologists physical status (ASA).^27^ The diagnoses of preoperative hypertension and diabetes were extracted from the medical records of the patients. Preoperative transient ischemic attack (TIA) or stroke was determined using the medical records of the patients and scores from neuroradiologists on cortical, subcortical and lacunar infarcts, based on the STRIVE criteria,^28^ as previously described by Kant and colleagues.^29^

### Delirium assessment

Delirium was defined according to the 5th edition of the Diagnostic and Statistical Manual of Mental Disorders (DSM-5) criteria.^1^ The Confusion Assessment Method for the Intensive Care Unit (CAM-ICU),^30^ the Nursing Delirium Screening Scale (Nu-DESC)^31^ and chart review^32^ were used to assess delirium by trained research staff. Patients were considered delirious in case of 2 or more cumulative points on the Nu-DESC and/or a positive CAM-ICU score and/or a chart review that showed clear descriptions of delirium (e.g., confused, agitated, drowsy, disorientated, delirious, receiving delirium-related antipsychotic therapy) that were confirmed by a consulted delirium expert. In case of uncertainty, a delirium expert was consulted to make the final decision on the diagnosis of the patient. If a patient was delirious, we additionally registered the duration of delirium (in days).

### Image processing

#### MRI scans

Imaging was performed on a 3T Achieva (Philips Medical Systems, Best, the Netherlands) scanner in Utrecht and on a 3T TrioTim (Siemens Healthineers, Erlangen, Germany) scanner in Berlin. A T1-weighted 3D Turbo Field Echo (TFE) structural image was made in Utrecht or a T1-weighted Magnetization Prepared Rapid Gradient Echo (MPRAGE) structural image was made in Berlin. A T2*-weighted gradient-echo – echoplanar imaging (GE-EPI) image was used for the resting-state blood-oxygen-level dependent (BOLD) fMRI (rs-fMRI) scan with a duration of approximately 8 minutes.^33^Preprocessing of the images was performed as previously described by Van Montfort and colleagues^33^ and in Supplemental Digital Content 1. Motion during fMRI measurements can induce bias^34–36^, therefore additional motion correction was conducted as reported previously^33^ and in Supplemental Digital Content 1.

#### Connectivity and network analysis

We selected 264 putative functional areas that cover all cortical and subcortical brain regions.^37^ All connectivity and network calculations were performed in MATLAB, version R2016b, using publicly available scripts and personalized scripts for the calculation of the MST metrics, as defined by Tewarie and colleagues.^38,39^ To estimate regional mean time series, voxel time series within each region were averaged. Using Pearson’s correlations, functional connectivity was subsequently calculated between all time series pairs, resulting in a 264×264 functional connectivity matrix for each patient.

Minimum spanning tree (MST) network backbones were extracted using Kruskal’s algorithm.^40^ The MST connections were all based on positive correlation values, thus avoiding the problematic interpretation of negative BOLD correlations.^38,39^ The MST can be considered as the backbone of the original network, connecting all regions without forming loops,^38–40^ which allows a relatively unbiased comparison with another network with the same number of regions and connections.^38,39^ Correlation values of the connectivity matrix were ranked and the highest value was included as the first MST connection using Kruskal’s algorithm.^40^ The second highest value was then added as an MST connection, etcetera until all 264 regions were connected. If adding a connection would result in a loop or triangle, this connection was discarded and the next value was evaluated.

Formally, a maximum spanning tree was thus constructed; the highest connectivity values were used to construct the MST as these connections were expected to reflect communication with minimal cost. To be consistent with previous literature using this approach, we refer to the minimum spanning tree or MST throughout this manuscript.

During delirium, altered global functional network connectivity strength, network efficiency, network organization and altered regional connectivity between the PCC and the DLPFC have been shown, therefore these outcomes were investigated in this study.^10,11,14^

#### Global functional connectivity strength

For each patient, global functional connectivity strength was calculated by averaging the connectivity values of all connections in the MST. This approach was chosen to maximize the signal-to-noise ratio of included connections within a standardized network backbone that is expected to reflect major information processing routes, to avoid problematic interpretation of negative BOLD correlations, and methodological bias of spurious connections or arbitrary thresholding in group comparisons of network topology.^38,39,41^

#### Network efficiency (MST diameter)

To assess network efficiency, the MST diameter was used. The MST diameter describes the number of edges connecting the most remote nodes in the MST and therefore gives an indication of the efficiency of global network organization.^38,39^ A low MST diameter indicates efficient information processing between remote brain regions.^10,38,39^

#### Network integration (MST leaf fraction)

To estimate the network integration, the MST leaf fraction was used. The MST leaf fraction describes the proportion of regions with a degree of one, i.e. regions that are connected to only one other region.^38,39^ A large MST leaf fraction indicates an integrated network topology.^38,39,42^

#### Functional connectivity between the PCC and the DLPFC (PCC-DLPFC-FC)

The PCC was defined as the region centered at coordinates (MNI x/y/z): –11/–56/16 (Power atlas region #77), −3/-49/13 (Power atlas region #78) and 11/-54/17 (Power atlas region #82).^14,37^ The DLPFC left was defined as the region centered at coordinates (MNI x/y/z): −42/38/21 (Power atlas region #167) and −34/55/4 (Power atlas region #176).^14,34^ The DLPFC right was defined as the region centered at coordinates (MNI x/y/z): 38/43/15 (Power atlas region #168) and 40/18/40 (Power atlas region #175).^14,34^ The connection between the PCC and the left or the right DLPFC was calculated for each patient using Pearson’s correlations between the mean time series of the regions.

### Statistical analysis

Statistical analyses were performed in R statistics (Version 3.5.1). Baseline characteristics were compared between patients who developed postoperative delirium and patients that did not, using the Chi-square test for categorical variables and independent samples t-test or Mann–Whitney U-test for continuous variables as appropriate.

We conducted generalized linear mixed models to analyze the change in global functional connectivity strength, MST diameter, MST leaf fraction and PCC-DLPFC-FC derived from the baseline and follow-up fMRI measurements in the same patients. ^43^ Confounders included age, sex, MMSE, surgical specialty, surgery duration and center. Selection of confounders was made based on clinical reasoning and knowledge obtained from literature. ^2,44^ As exploratory analysis, we performed Spearman’s correlation analyses to investigate the association between the MST measures (global functional connectivity strength, MST diameter and MST leaf fraction) and duration of the delirium (in patients that developed postoperative delirium only). Generalized mixed models were also used to compare change in TMT score (A or B) between the delirium and the non-delirium group, and for a posthoc analysis for fMRI outcome measures with a significant change between the delirium and the non-delirium group three months postoperatively compared to preoperatively. In this analysis, we evaluated whether this change in fMRI outcome was related to change in cognitive performance as measured with TMT scores. The group that decreased on the fMRI outcome measure was compared with regard to the TMT change three months postoperatively compared to preoperatively to the group that increased or remained stable on the fMRI outcome, using a generalized linear mixed model. Two-sided tests were used for all analyses and a p-value below 0.05 was considered statistically significant. More detailed information on the modeling procedure is available in Supplemental Digital Content 2.

## Results

### Demographics

In this study, the total eligible cohort consisted of 554 patients that performed baseline measurements. In total, 246 patients with sufficient quality of the pre- and or postoperative fMRI scan and available postoperative data, were included in this study (Supplemental Digital Content 3). The included patients were generally more often from center Utrecht, younger, had less comorbidities and scored higher on the TMT B than excluded patients (Supplemental Digital Content 4). Of the 246 included patients, 38 (16%) developed delirium within the first seven postoperative days. Patients who developed delirium were generally older, had a longer duration of surgery and had a longer hospital stay (Table 1).

**Table 1.**
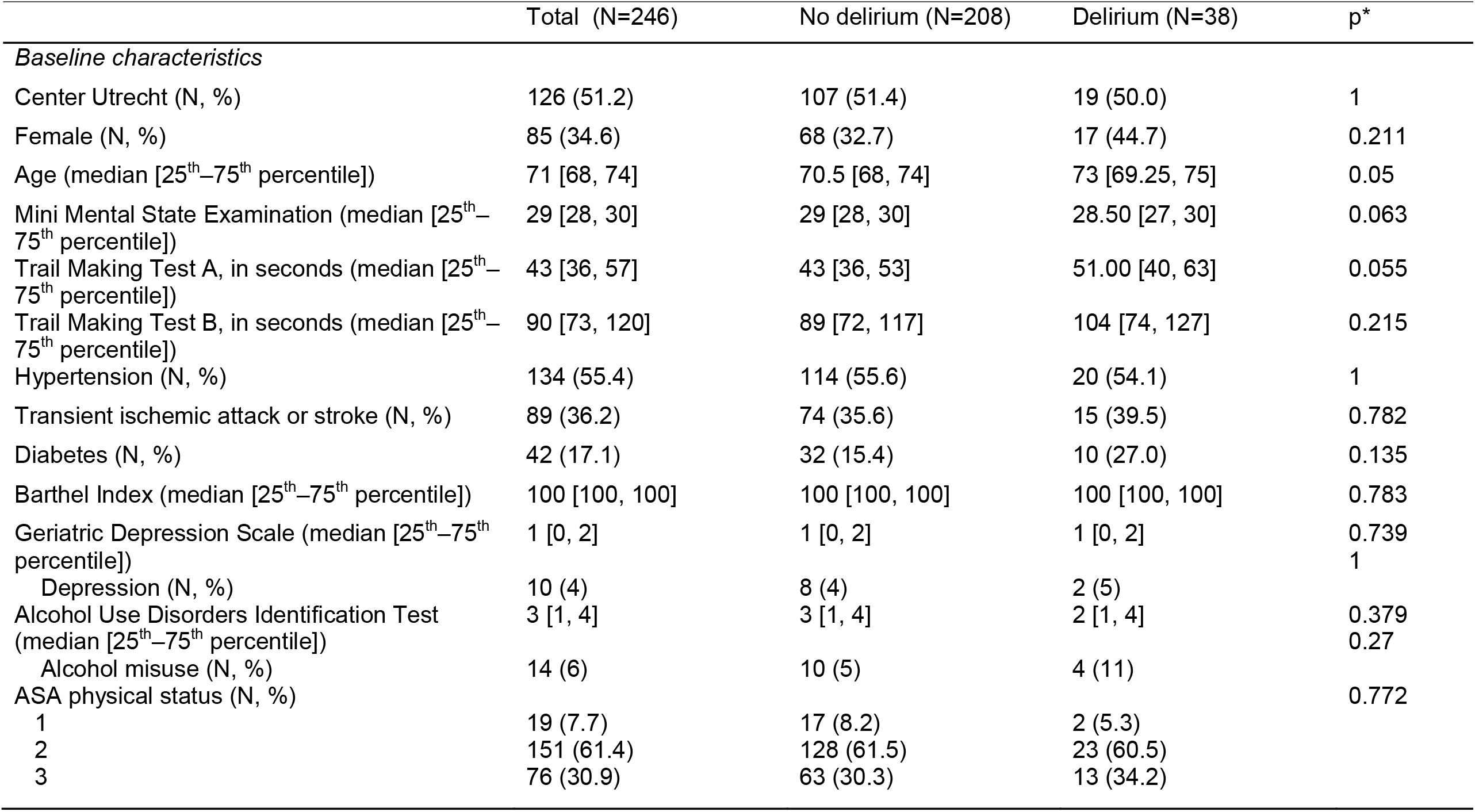
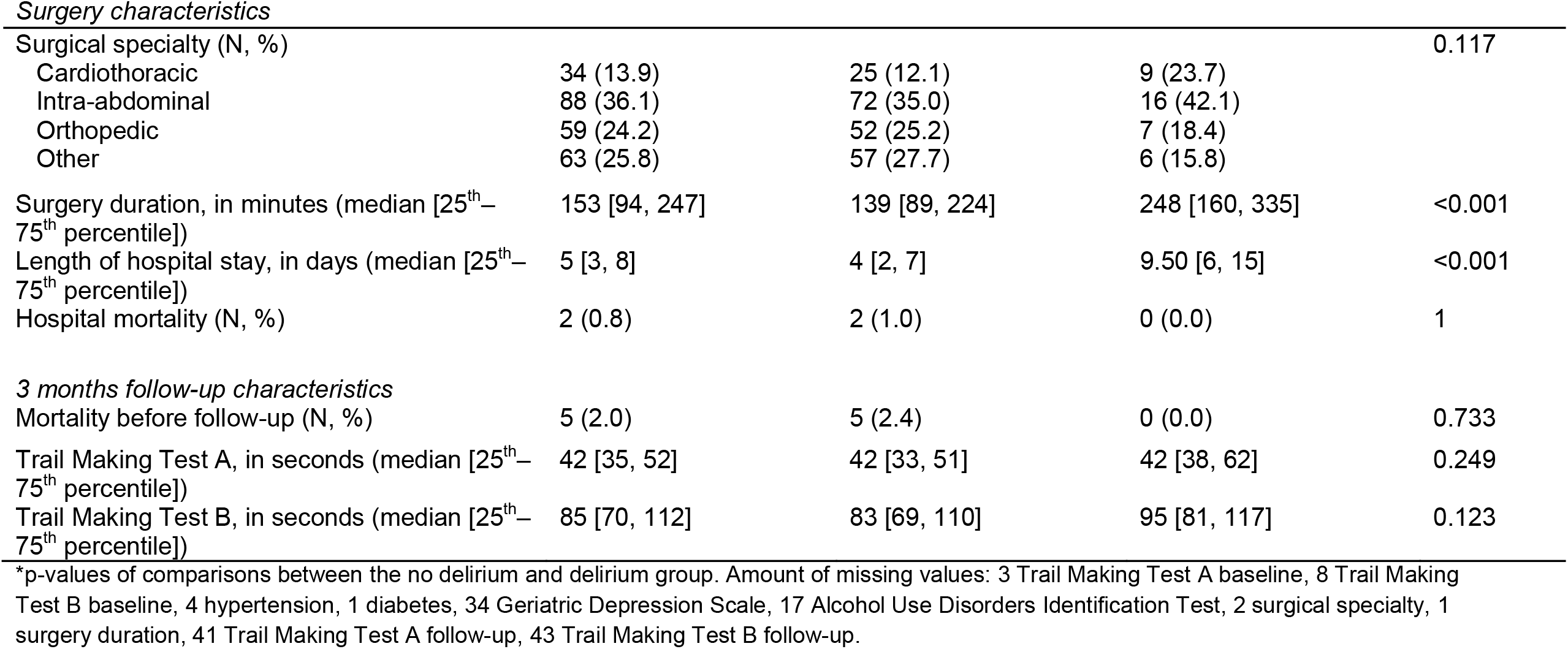
Characteristics of the total included sample.

|  | Total (N=246) | No delirium (N=208) | Delirium (N=38) | p* |
| --- | --- | --- | --- | --- |
| *Baseline characteristics* |  |  |  |  |
| Center Utrecht (N, %) | 126 (51.2) | 107 (51.4) | 19 (50.0) | 1 |
| Female (N, %) | 85 (34.6) | 68 (32.7) | 17 (44.7) | 0.211 |
| Age (median [25<sup>th</sup>–75<sup>th</sup> percentile]) | 71 [68, 74] | 70.5 [68, 74] | 73 [69.25, 75] | 0.05 |
| Mini Mental State Examination (median [25<sup>th</sup>–75<sup>th</sup> percentile]) | 29 [28, 30] | 29 [28, 30] | 28.50 [27, 30] | 0.063 |
| Trail Making Test A, in seconds (median [25<sup>th</sup>–75<sup>th</sup> percentile]) | 43 [36, 57] | 43 [36, 53] | 51.00 [40, 63] | 0.055 |
| Trail Making Test B, in seconds (median [25<sup>th</sup>–75<sup>th</sup> percentile]) | 90 [73, 120] | 89 [72, 117] | 104 [74, 127] | 0.215 |
| Hypertension (N, %) | 134 (55.4) | 114 (55.6) | 20 (54.1) | 1 |
| Transient ischemic attack or stroke (N, %) | 89 (36.2) | 74 (35.6) | 15 (39.5) | 0.782 |
| Diabetes (N, %) | 42 (17.1) | 32 (15.4) | 10 (27.0) | 0.135 |
| Barthel Index (median [25<sup>th</sup>–75<sup>th</sup> percentile]) | 100 [100, 100] | 100 [100, 100] | 100 [100, 100] | 0.783 |
| Geriatric Depression Scale (median [25<sup>th</sup>–75<sup>th</sup> percentile]) | 1 [0, 2] | 1 [0, 2] | 1 [0, 2] | 0.739 |
| Depression (N, %) | 10 (4) | 8 (4) | 2 (5) | 1 |
| Alcohol Use Disorders Identification Test (median [25<sup>th</sup>–75<sup>th</sup> percentile]) | 3 [1, 4] | 3 [1, 4] | 2 [1, 4] | 0.379 |
| Alcohol misuse (N, %) | 14 (6) | 10 (5) | 4 (11) | 0.27 |
| ASA physical status (N, %) |  |  |  | 0.772 |
| 1 | 19 (7.7) | 17 (8.2) | 2 (5.3) |  |
| 2 | 151 (61.4) | 128 (61.5) | 23 (60.5) |  |
| 3 | 76 (30.9) | 63 (30.3) | 13 (34.2) |  |
| *Surgery characteristics* |  |  |  |  |
| Surgical specialty (N, %) |  |  |  | 0.117 |
| Cardiothoracic | 34 (13.9) | 25 (12.1) | 9 (23.7) |  |
| Intra-abdominal | 88 (36.1) | 72 (35.0) | 16 (42.1) |  |
| Orthopedic | 59 (24.2) | 52 (25.2) | 7 (18.4) |  |
| Other | 63 (25.8) | 57 (27.7) | 6 (15.8) |  |
| Surgery duration, in minutes (median [25<sup>th</sup>–75<sup>th</sup> percentile]) | 153 [94, 247] | 139 [89, 224] | 248 [160, 335] | <0.001 |
| Length of hospital stay, in days (median [25<sup>th</sup>–75<sup>th</sup> percentile]) | 5 [3, 8] | 4 [2, 7] | 9.50 [6, 15] | <0.001 |
| Hospital mortality (N, %) | 2 (0.8) | 2 (1.0) | 0 (0.0) | 1 |
| *3 months follow-up characteristics* |  |  |  |  |
| Mortality before follow-up (N, %) | 5 (2.0) | 5 (2.4) | 0 (0.0) | 0.733 |
| Trail Making Test A, in seconds (median [25<sup>th</sup>–75<sup>th</sup> percentile]) | 42 [35, 52] | 42 [33, 51] | 42 [38, 62] | 0.249 |
| Trail Making Test B, in seconds (median [25<sup>th</sup>–75<sup>th</sup> percentile]) | 85 [70, 112] | 83 [69, 110] | 95 [81, 117] | 0.123 |

\*p-values of comparisons between the no delirium and delirium group. Amount of missing values: 3 Trail Making Test A baseline, 8 Trail Making Test B baseline, 4 hypertension, 1 diabetes, 34 Geriatric Depression Scale, 17 Alcohol Use Disorders Identification Test, 2 surgical specialty, 1 surgery duration, 41 Trail Making Test A follow-up, 43 Trail Making Test B follow-up.

fMRI data of sufficient quality was available for 216 patients at baseline and for 160 patients at three months follow-up. Of these patients, 130 had fMRI data of sufficient quality available at both measurements (Supplemental Digital Content 5). Patients that had two fMRI scans of sufficient quality available, were generally more often from center Utrecht, younger and had a better performance on TMT B. Delirium was diagnosed in 18 patients (14%) in this group.

The evaluated fMRI outcomes appeared stable over the used timespan, tested in a group of non-surgical controls (Supplemental Digital Content 6 and 7).

### Change in TMT scores

TMT values at baseline and follow-up are described in Table 1. The delirium and the non-delirium group did not differ in TMT scores at three months follow-up compared to preoperatively (TMT A (β = 0.63, 95% CI= −5.27 - 6.52, p = 0.835), TMT B (β = −0.43, 95% CI= −12.39 −11.54, p = 0.944 and TMT ratio, β=0.06, 95% CI= −0.24 −0.36, p=0.694).

### Global functional connectivity strength

Patients who developed postoperative delirium did not differ from patients who remained delirium-free regarding global functional connectivity strength prior to surgery (β = 0.006, 95% CI = −0.003 – 0.016, p = 0.165) (for mean values, see Supplemental Digital Content 7). Compared to preoperatively, global functional connectivity strength increased three months postoperatively in the total study population (β = 0.006, 95% CI = 0.000 – 0.012, p = 0.021). However, a significant interaction between delirium and measurement (i.e. baseline and follow-up) was found showing a decline in global functional connectivity strength for patients who developed delirium, relative to an increase in global functional connectivity strength in patients who remained delirium-free (β = −0.014, 95% CI = −0.028 – −0.008, p = 0.026) (Figure 1). In the group of 130 patients with both fMRI scans available at baseline and follow-up, no change in global functional connectivity strength between the preoperative and follow-up measurement was observed (β= 0.004, 95% CI= −0.000-0.010, p=0.063). However, a decline in global functional connectivity strength was still present for patients who developed delirium (β= −0.017, 95% CI= −0.032--0.002, p=0.021). No significant correlations were found between duration of delirium and global functional connectivity strength at three months (ρ = −0.067, p = 0.787), or change in global functional connectivity strength (ρ = −0.051, p = 0.852).

**Figure 1.**
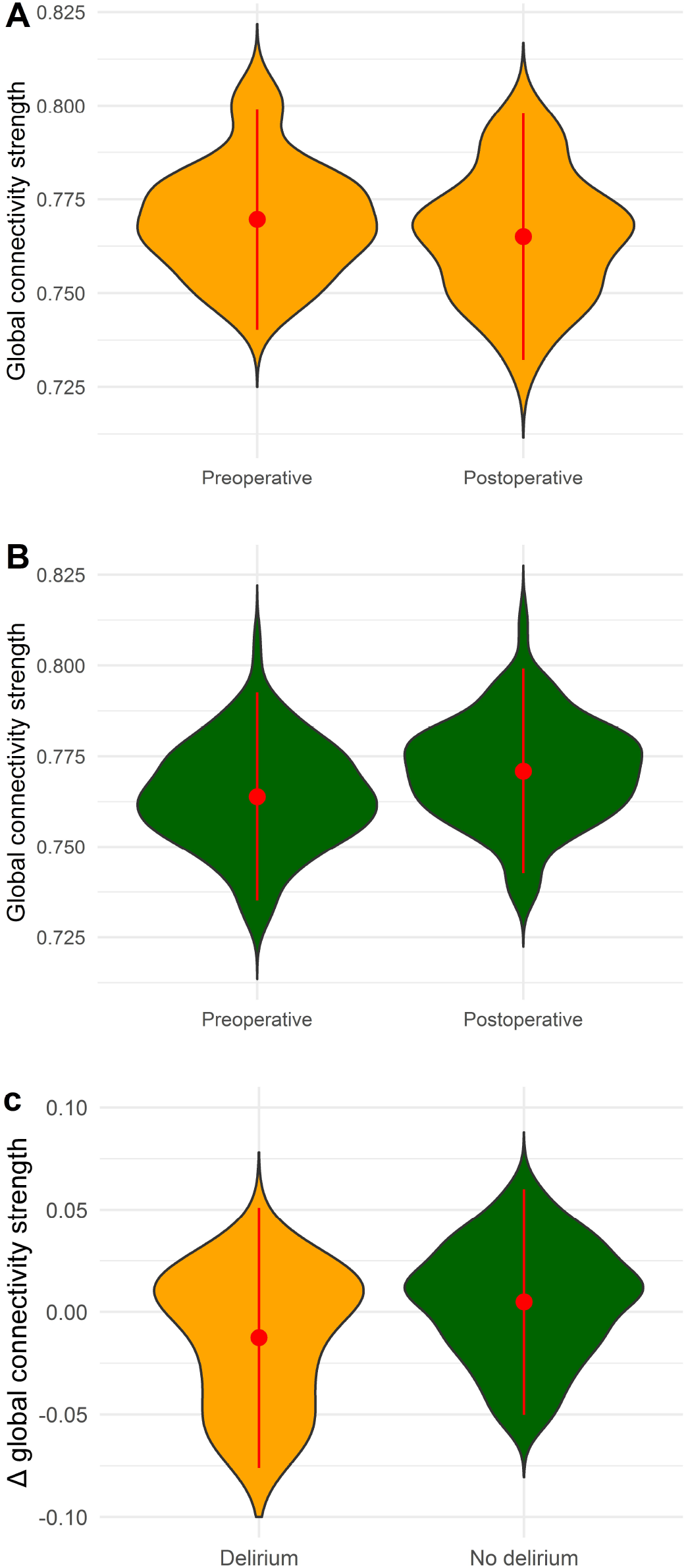
Global functional connectivity strength preoperatively versus three months postoperatively for the delirium (panel A; N=38) and non-delirium group (panel B; N=208). Global functional connectivity strength was evaluated at baseline (preoperatively) and at three months follow-up, in an elderly population undergoing major, elective surgery. Global functional connectivity strength was significantly decreased in patients that developed postoperative delirium three months after surgery, but was increased in patients that did not develop postoperative delirium. Panel C presents the difference in global functional connectivity strength between the preoperative and postoperative measurements for delirious (N=19) and non-delirious patients (N=121) separately. Difference in global functional connectivity strength could only be obtained for patients with complete data (N=130).

### Global functional connectivity strength & change in TMT scores

In the group of 130 patients with sufficient quality fMRI scans available, 52 patients (40%) showed a decrease in global functional connectivity strength three months postoperatively compared to preoperatively. Patients with decreased global functional connectivity strength at follow-up (with 15.4% incidence of postoperative delirium) had significantly declined in TMT B score (β = 11.04, 95% CI= 0.85-21.2, p = 0.034) compared to patients with equal or increased global functional connectivity strength at follow-up (10.3% incidence of postoperative delirium) (Figure 2). No change in TMT A and TMT ratio was found between the preoperative and postoperative fMRI measurement among patients with decreased functional connectivity strength(TMT A; β = 3.06, 95% CI= −2.83-8.96, p = 0.31, TMT ratio; β = 0.07, 95% CI= −0.21-0.36 p = 0.598) (Figure 3).

**Figure 2.**
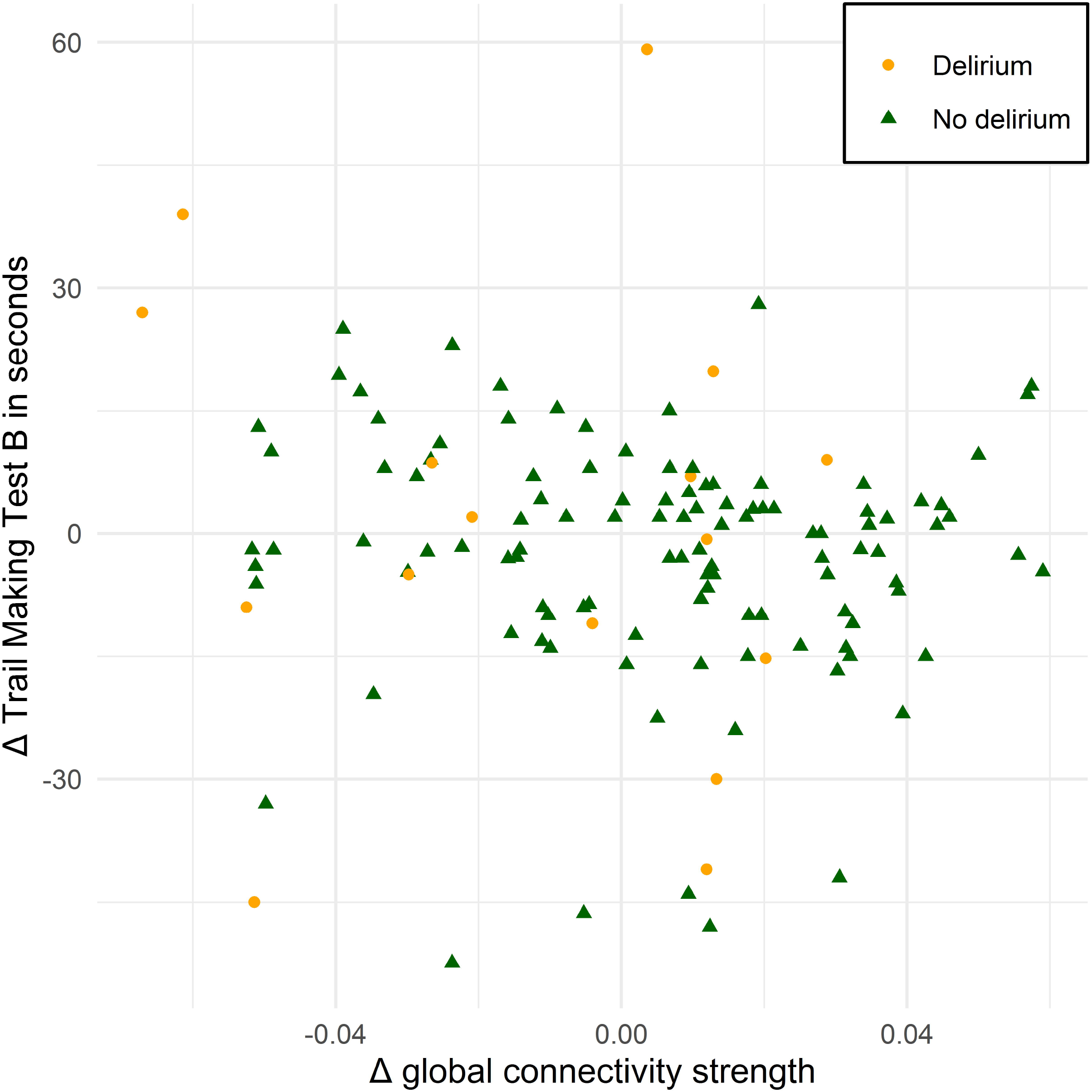
Scatter plot of change in global functional connectivity strength between the preoperative and postoperative fMRI at three months after surgery against change in Trail Making Test B scores measured at the same time points as the fMRI (ρ=-0.230, p=0.010). The lower the score (or large decrease in score), the better the performance. No interaction with delirium was found. fMRI = functional magnetic resonance imaging.

**Figure 3.**
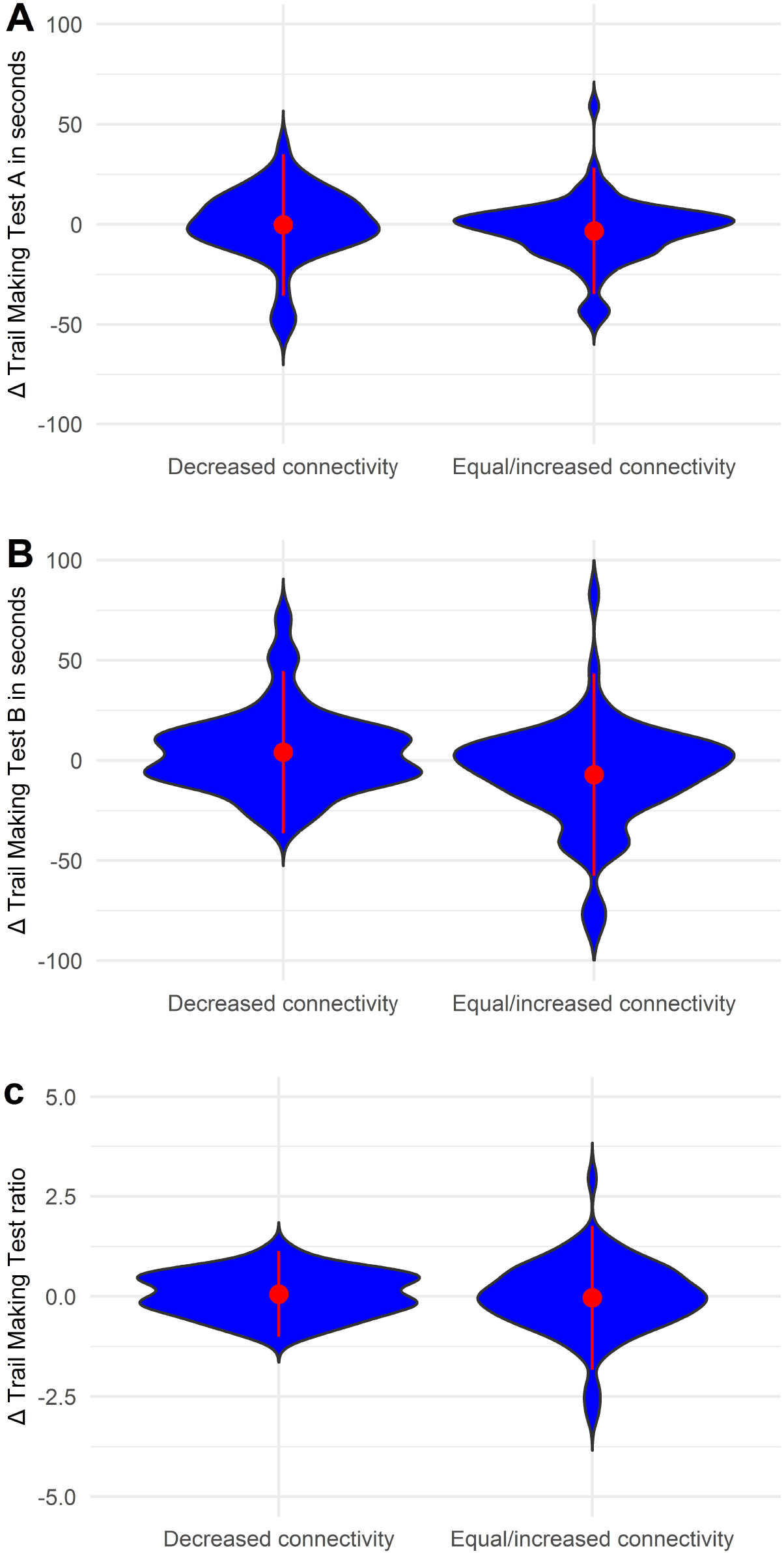
Trail making test scores preoperatively versus three months postoperatively for the group that decreased in global functional connectivity strength and the group that increased or remained equal in global functional connectivity strength. Trail Making Test A and B scores are reaction times in seconds, the lower the scores, the better the performance. Change in Trail Making Test A scores were not different among patients with decreased global functional connectivity strength compared to patients with increased/equal global functional connectivity strength (A). The group that decreased in global functional connectivity strength significantly declined in Trail Making Test B performance (B). Normally, a learning effect occurs when the task is repeated, which can be observed in the group with an equal or increased global functional connectivity strength. No differences were observed between both groups for Trail Making Test ratio (Trail Making Test B / Trail Making Test A) (C).

### Functional network efficiency (MST diameter)

No differences were found between delirium and non-delirium patients on MST diameter at baseline (β = 0.004, 95% CI = −0.004 – 0.012, p = 0.339), (see Supplemental Digital Content 8). Compared to preoperatively, no significant change was found in MST diameter three months postoperatively for the total study population (β = 0.000, 95% CI = −0.004 – 0.005, p = 0.871). In addition, no interaction was found for delirium and non-delirium patients at three months postoperatively compared to preoperatively in MST diameter (β = 0.000, 95% CI = −0.013 – 0.013, p = 0.976) (Supplemental Digital Content 9). No significant correlations were found between delirium duration and MST diameter at three months follow-up (ρ = 0.029, p = 0.907) or change in MST diameter (ρ = 0.234, p = 0.383). Results were similar in the subgroup of patients with both fMRI scans available (data not shown).

### Functional network integration (MST leaf fraction)

No differences were found between delirium and non-delirium patients on MST leaf fraction at baseline (β = 0.000, 95% CI = −0.009 – 0.008, p = 0.843), (see Supplemental Digital Content 8). Compared to preoperatively, no significant change was found in MST leaf fraction three months postoperatively for the total study population (β = −0.003, 95% CI = −0.008 – 0.001, p = 0.134). In addition, no interaction was found for delirium and non-delirium patients at three months postoperatively compared to preoperatively in MST leaf fraction (β = 0.002, 95% CI = −0.001 – −0.001, p = 0.771) (Supplemental Digital Content 9). No significant correlations were found between delirium duration and MST leaf fraction at three months follow-up (ρ = −0.036, p = 0.883) or change in MST leaf fraction (ρ = −0.179, p = 0.506). Again, similar results were found in patients with both fMRIs (data not shown).

### PCC-DLPFC-FC

No differences were found between delirium and non-delirium patients on PCC-DLPFC-FC at baseline (left: β = −0.018, 95% CI = −0.059 – 0.024, p = 0.397, right: β = −0.001, CI = −0.044 – 0.043, p = 0.979), (Supplemental Digital Content 8). Compared to preoperatively, no significant change was found in PCC-DLPFC-FC three months postoperatively for the total study population (left: β = 0.022, 95% CI = −0.002 – 0.047, p = 0.007, right: β = −0.005, 95% CI = −0.029 – 0.019, p = 0.669). In addition, no interaction was found for delirium and non-delirium patients at three months postoperatively compared to preoperatively in PCC-DLPFC-FC (left: β = 0.010, 95% CI = −0.002 – 0.047, p = 0.757, right: β = 0.002, 95% CI = −0.029 – 0.019, p = 0.858) (Supplemental Digital Content 9).

## Discussion

In this longitudinal study, we investigated the impact of major surgery and postoperative delirium on fMRI functional brain networks in an elderly population. Three months after surgery and relative to the preoperative measurements, global functional connectivity strength was decreased in patients who had suffered postoperative delirium, but increased in patients who remained delirium-free. Patients who showed decreased global functional connectivity strength at follow-up had a decline in executive function compared to patients with an increased global functional connectivity strength, irrespective of delirium. This study is the first to empirically evaluate the changes in the functional brain network preoperatively compared to three months postoperatively, in relation to the presence or absence of postoperative delirium, and to link these changes in the functional brain network to postoperative cognitive performance.

No baseline differences in functional brain networks were found between patients who developed delirium postoperatively and those who remained delirium-free, which is in line with previous studies showing that functional brain network characteristics that are altered during delirium do not necessarily reflect delirium vulnerability.^10,33^ The onset of delirium seems therefore to reflect new functional network impairments. However, the dynamic nature of the functional brain network, as well as the complex interaction between brain network structure and function, might imply that other network characteristics predispose patients to delirium than those observed during the syndrome itself.^6^

In a previous study, we found that global functional connectivity strength was lower in patients one week after delirium compared to healthy controls,^10^ but no fMRI data were available before the onset of delirium in that investigation. The findings presented in the current study show that the decrease of global functional connectivity strength is even found three months after remission of the syndrome. The decrease was associated with deteriorated executive function, as measured with the TMT B. As decreased global functional connectivity strength is also observed in patients with severe cognitive impairment and dementia,^12,13,17^ we speculate that the lasting decrease in global functional connectivity strength could be related to poor cognitive outcomes in some patients after recovery from surgery, such as long-term cognitive impairment. As we found no decline in TMT B test scores at follow-up between patients with and without postoperative delirium, we could not test if delirium-related cognitive decline was mediated by decreased global functional connectivity strength.

The absence of long-term TMT impairment in the delirium group may reflect a lack of impact of delirium on long-term executive functioning, but it may also be explained by several other factors. It could be that the patients that had more severe cognitive problems were lost to follow-up, as they were unable to come to the hospital for the follow-up measurements. Furthermore, although delirium is a risk factor for long-term cognitive impairment, not all patients will develop long-term cognitive problems. We may have included a relatively healthy group of elective surgery patients, due to an extensive study protocol and strict motion correction.^18^ This group may, in general, have had higher cognitive reserves and strong brain plasticity, possibly resulting in the ability to cognitively recover from delirium.^45^

An interesting observation is that major surgery was associated with an increase in global functional connectivity three months postoperatively compared to preoperatively. However, it is known that surgery alone may also increase the risk of long-term cognitive dysfunction.^46^ The interpretation of general alterations in connectivity strength is not straightforward. A simulation study of activity-dependent neural degeneration indicated that increased functional connectivity strength may reflect high neural activity levels, which may result in neural damage and could therefore be considered as an early state of vulnerability.^47^ As major surgery may result in neuroinflammation and reduced oxygen supply, the increase of global functional connectivity strength in the majority of patients without postoperative delirium may be a reflection of the resilient brain response to the complex interaction of these (and possibly other) factors.^47,48^

At three months follow-up, we found that postoperative delirium was not associated with changes in functional network efficiency, functional network integration or regional functional connectivity between the posterior cingulate cortex and the dorsolateral prefrontal cortex. This finding is consistent with the previously mentioned study on functional brain networks that showed that seven days after delirium was clinically resolved, there were no lasting changes in functional network efficiency or network integration.^10^ Alterations in functional network efficiency and functional network integration seem thus to be related to the clinical syndrome of delirium and may recover when delirium resolves.

An important strength of this study is that we were able to obtain measurements before the onset of delirium by studying surgical patients preoperatively. In addition, robust methods were used and a large number of patients was included in this multicenter study. However, there are also important limitations. Due to rigorous motion correction, a considerable part of our study population had to be excluded, which may have resulted in selection of patients who are less vulnerable for delirium. Furthermore, we specifically focused on the TMT in this study, because decreased TMT test scores have previously been associated with delirium severity and also with decreased global functional connectivity strength (measured with EEG) in dementia with Lewy bodies.^16,21^ However, long-term cognitive impairment after delirium based on functional network changes may additionally be reflected in other cognitive tasks. Another limitation is that information on medication use of the patients was not stored. We can therefore not exclude possible drug-related effects on the fMRI measurements. Nevertheless, all patients were non-hospitalized during the fMRI measurements. Further, we focused on fMRI network characteristics that are altered during delirium, and did not evaluate the relationship between delirium risk factors and all possible fMRI network changes or used seed-based analyses to focus on unexplored regional connections that are altered in patients after surgery or delirium. It could therefore be that functional brain impairments related to surgery or delirium are represented in other fMRI outcomes.

### Conclusion

Delirium seems to result in a decrease in global functional connectivity strength, three months after the syndrome was resolved. Decreased global functional connectivity strength was associated (but not necessarily clinically relevant) with cognitive decline, irrespective of postoperative delirium. Notably, due to the exclusion of patients with fMRI scans of insufficient quality, which were generally older, had more comorbidities and scored worse on the TMT B, our results could have been influenced by selection effects. Still, our study may provide new insights into the biological substrate of long-term brain changes after surgery and delirium.

## Data Availability

All data produced in the present study are available upon reasonable request to the authors

## Acknowledgment

We would like to especially thank Noam Rubin, Raoul Lieben and Rutger van de Leur for the technical support with the fMRI analyses and Jacqueline Vromen and Wietze Pasma for the support with data management.

## Abbreviations

ASA: American Society of Anesthesiologists physical status
AIC: Akaike’s information criterion
AUDIT: Alcohol Use Disorders Identification Test
BioCog: Biomarker Development for Postoperative Cognitive Impairment in the Elderly
CAM-ICU: Confusion Assessment Method for the Intensive Care Unit
CEN: Central executive network
DLPFC: Dorsolateral prefrontal cortex
DMN: Default mode network
DSM-5: Diagnostic and Statistical Manual of Mental Disorders
fMRI: Functional magnetic resonance imaging
GDS: Geriatric Depression Scale
GE-EPI: Gradient-echo – echoplanar imaging
MMSE: Mini Mental State Examination
MNI: Montreal Neurological Institute
MPRAGE: Magnetization Prepared Rapid Gradient Echo
MST: Minimum spanning tree
Nu-DESC: Nursing Delirium Screening Scale
PCC: Posterior cingulate cortex
PCC-DLPFC-FC: Functional connectivity between the PCC and the DLPFC
POD+: Patients with postoperative delirium
POD-: Patients without postoperative delirium
REML: Restricted maximum likelihood estimation
Rs-fMRI: Resting-state functional magnetic resonance imaging
TFE: Turbo Field Echo
TIA: Transient ischemic attack
TMT: Trail Making Test
TMT-A: Trail Making Test section A
TMT-B: Trail Making Test section B
UMC: University Medical Center

