## Supplementary material for "Functional brain network and trail making test changes after major surgery and delirium"

Supplemental Digital Content 1. Detailed information on preprocessing and motion correction.

Preprocessing and motion correction

Preprocessing of the images was performed using the FMRIB’s Software Library (FSL) ^1–3^. The brain was automatically extracted from the T1-weighted scan ^4^. The time series were corrected for motion with MCFLIRT ^5,6^. Motion during fMRI measurements can induce bias, therefore additional motion correction is necessary ^7–11^. Volumes that exceeded the threshold of 0.2 mm framewise displacement ^12^ were removed and a regression analysis with 36 motion components was done. The motion components were: three voxel-wise displacement parameters and their white matter, cerebrospinal fluid, global time courses, and the quadrates, temporal derivatives and quadrates of the derivatives of these six parameters ^10^. The average time series from the cerebral spinal fluid, the white matter and grey matter intensities were defined after tissue segmentation with the FMRIB's Automated Segmentation Tool (FAST) ^13^. A band-pass filter (0.01 – 0.08 Hz) was applied ^10^.

The functional scan was registered to the high-resolution anatomical image using rigid registration. The anatomical scan was subsequently registered to the Montreal Neurological Institute (MNI) 152 T1-weighted 2 mm image in standard space with affine registration. Functional scans were slice-time corrected and spatially smoothed to reduce noise (5 mm full-width-half-maximum). To ensure stabilized magnetization, the first 15 volumes were deleted. If the remaining data was less than 240 seconds, the patient was excluded from further analysis ^14^.

Supplemental Digital Content 2. Statistical analysis

No formal statistical power calculation was conducted prior to this study. The sample size was based on available data. Descriptive data was reported as frequencies with percentage for categorical data and means with standard deviations (SD) or median with 25th and 75th percentile (interquartile range, IQR) for continuous data, as appropriate. Normality for continuous variables was examined by visually inspecting histograms and normal quantile–quantile plots. Baseline characteristics were compared between patients who developed postoperative delirium and patients that did not, using the Chi-square test for categorical variables and independent samples t-test or Mann–Whitney U-test for continuous variables as appropriate. Surgical specialty was categorized in cardiothoracic, intra-abdominal, orthopedic and other. Surgery duration was analyzed per minute and duration of delirium in days.

We conducted generalized linear mixed models to analyze the change in global functional connectivity strength, MST diameter, MST leaf fraction and PCC-DLPFC-FC derived from the baseline and follow-up fMRI measurements (i.e. three months after surgery) in the same patients. In addition, we investigated whether these changes in network and connectivity measures were different between patients that developed postoperative delirium and patients that did not. Univariable analyses were performed with timing of the measurement (preoperative or three months after surgery) and delirium included in the fixed part of the model and ‘patient’ was added as a random intercept. To assess whether global functional connectivity strength, MST leaf fraction, and MST diameter varied over the two measurements (preoperatively and three months postoperatively) among patients who developed postoperative delirium and patients that did not, an interaction term was added between delirium and time in the fixed part of the model. Patients with only one measurement available were also included in the model. ^1^ Thereafter, we conducted multivariable analyses by adding *a priori* selected confounders to the fixed part of the model. These included age, sex, MMSE, surgical specialty, surgery duration and center. Selection of confounders was made based on clinical reasoning and knowledge obtained from literature.^2,3^ Models were compared based on the Akaike’s information criterion (AIC). Restricted maximum likelihood estimation (REML) was used to generate unbiased variance estimates for the final models. Estimates are expressed as linear regression coefficients (β) with 95% confidence intervals (95% CI). To explore the effect of potential follow-up bias, we repeated the complete analytical approach described above for patients with complete data on both fMRI measurements as a sensitivity analysis.

As exploratory analysis, we performed Spearman’s correlation analyses to investigate the association between the MST measures (global functional connectivity strength, MST diameter and MST leaf fraction) and duration of the delirium (in patients that developed postoperative delirium only). Change in TMT score (A or B) was compared between the delirium and the non-delirium group using a generalized linear mixed model. The model included an interaction term between delirium and timing of the measurement (i.e. baseline or follow-up measurement) in the fixed part and ‘patient’ was added as an random intercept. A posthoc analysis was conducted for fMRI outcome measures with a significant change between the delirium and the non-delirium group three months postoperatively compared to preoperatively. In this analysis, we evaluated whether this change in fMRI outcome was related to change in cognitive performance as measured with TMT scores. The group that decreased on the fMRI outcome measure was compared with regard to the TMT change three months postoperatively compared to preoperatively to the group that increased or remained stable on the fMRI outcome, using a generalized linear mixed model. Again, an interaction term between decline in fMRI outcome and measurement (i.e. baseline or follow-up measurement) was included with ‘patient’ as a random intercept. Two-sided tests were used for all analyses and a p-value below 0.05 was considered statistically significant.

Supplemental Digital Content 3. Flowchart of the inclusion of subjects in this study.

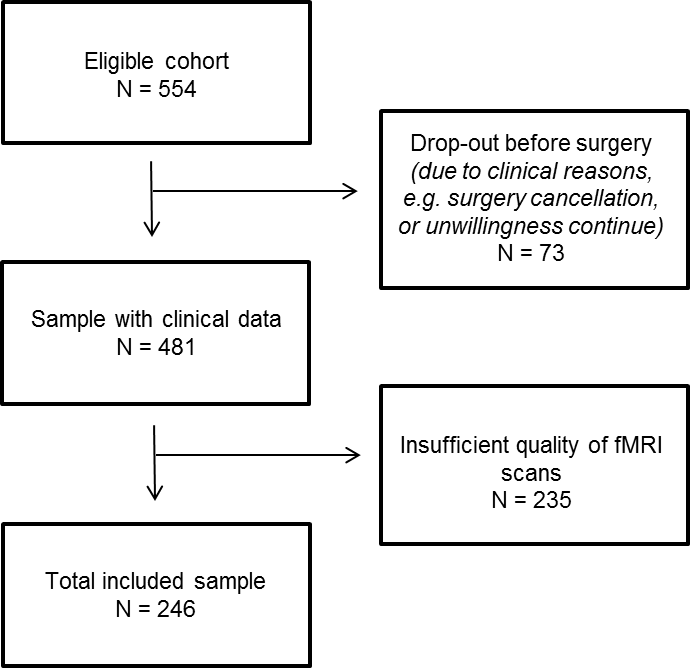

Supplemental Digital Content 4. Characteristics of the total sample with available clinical data and the included sample.

|  | Total sample with clinical data (N=481) | Included (N=246) | Excluded (N=235) | p |
| --- | --- | --- | --- | --- |
| *Baseline characteristics* | | | | |
| Center Utrecht (N, %) | 176 (36.7) | 126 (51.2) | 50 (21.4) | <0.001 |
| Female (N, %) | 186 (39.0) | 85 (34.6) | 101 (43.7) | 0.050 |
| Age (median [25^th^–75^th^ percentile]) | 72 [68, 75] | 71 [68, 74] | 72 [69, 76] | 0.026 |
| Mini Mental State Examination (median [25^th^–75^th^ percentile]) | 29 [28, 30] | 29 [28, 30] | 29 [27, 30] | 0.001 |
| Trail Making Test A, in seconds (median [25^th^–75^th^ percentile]) | 45 [37, 58] | 43 [36, 57] | 47 [37, 61] | 0.064 |
| Trail Making Test B, in seconds (median [25^th^–75^th^ percentile]) | 100 [79, 131] | 90 [73, 120] | 111 [92, 149] | <0.001 |
| Hypertension (N, %) | 289 (62.0) | 134 (55.4) | 155 (69.2) | 0.003 |
| Transient ischemic attack or stroke (N, %) | 172 (35.8) | 89 (36.2) | 83 (35.3) | 0.919 |
| Diabetes (N, %) | 105 (22.4) | 42 (17.1) | 63 (28.1) | 0.006 |
| Barthel Index (median [25^th^–75^th^ percentile]) | 100 [100, 100] | 100 [100, 100] | 100 [100, 100] | 0.065 |
| Geriatric Depression Scale (median [25^th^–75^th^ percentile])  Depression (N, %) | 1 [0, 2]  22 (5.6) | 1 [0, 2]  10 (4.7) | 1 [0, 3]  12 (6.6) | 0.017  0.565 |
| Alcohol Use Disorders Identification Test (median [25^th^–75^th^ percentile])  Alcohol misuse (N, %) | 2 [1, 4]  24 (5.5) | 3 [1, 4]  14 (6.1) | 2 [0, 4]  10 (4.8) | 0.017  0.698 |
| ASA physical status (N, %)  1  2  3 | 26 (5.5)  284 (60.3)  161 (34.2) | 19 (7.7)  151 (61.4)  76 (30.9) | 7 (3.1)  133 (59.1)  85 (37.8) | 0.044 |
| Surgical specialty (N, %)  Cardiothoracic  Intra-abdominal  Orthopedic  Other | 44 (9.4)  155 (33.2)  136 (29.1)  132 (28.3) | 34 (13.9)  88 (36.1)  59 (24.2)  63 (25.8) | 10 (4.5)  67 (30.0)  77 (34.5)  69 (30.9) | 0.001 |
| Surgery duration, in minutes (median [25^th^–75^th^ percentile]) | 143 [89, 228] | 153 [94, 247] | 135 [82, 206] | 0.024 |
| Length of hospital stay, in days (median [25^th^–75^th^ percentile]) | 5 [3, 9] | 5 [3, 8] | 6 [3, 9] | 0.171 |
| Delirium (N, %) | 75 (16.3) | 38 (15.4) | 37 (17.3) | 0.684 |
| Hospital mortality (N, %) | 5 (1.1) | 2 (0.8) | 3 (1.4) | 0.900 |
| *3 months follow-up characteristics* |  |  |  |  |
| Mortality before follow-up (N, %) | 18 (3.9) | 5 (2.0) | 13 (5.9) | 0.054 |
| Trail Making Test A, in seconds (median [25^th^–75^th^ percentile]) | 42 [35, 53] | 42 [35, 52] | 45 [35, 58] | 0.240 |
| Trail Making Test B, in seconds (median [25^th^–75^th^ percentile]) | 93 [74, 119] | 85 [70, 112] | 100 [80, 131] | <0.001 |

*p-values of comparisons between the included and the excluded group.

Supplemental Digital Content 5. Characteristics of the subjects that had an fMRI scan of sufficient quality available baseline and follow-up or at one of the two timepoints.

|  | Total included sample (N=246) | Baseline + follow up fMRI (N=130) | Baseline fMRI only  (N=86) | Follow up fMRI only (N=30) | p |
| --- | --- | --- | --- | --- | --- |
| *Baseline characteristics* | | | | | |
| Center Utrecht (N, %) | 126 (51.2) | 84 (64.6) | 28 (32.6) | 14 (46.7) | <0.001 |
| Female (N, %) | 85 (34.6) | 48 (36.9) | 32 (37.2) | 5 (16.7) | 0.089 |
| Age (median [25^th^–75^th^ percentile]) | 71 [68, 74] | 70 [67, 74] | 72 [68, 74] | 73 [70, 76] | 0.015 |
| Mini Mental State Examination (median [25^th^–75^th^ percentile]) | 29 [28, 30] | 29 [28, 30] | 29 [28, 30] | 29 [28, 30] | 0.230 |
| Trail Making Test A, in seconds (median [25^th^–75^th^ percentile]) | 43 [36, 57] | 42 [36, 53] | 45 [36, 60] | 49 [40, 60] | 0.129 |
| Trail Making Test B, in seconds (median [25^th^–75^th^ percentile]) | 90 [73, 120] | 82 [70, 105] | 105 [81, 133] | 91 [76, 108] | <0.001 |
| Hypertension (N, %) | 134 (55.4) | 64 (50.0) | 49 (58.3) | 21 (70.0) | 0.111 |
| Transient ischemic attack or stroke (N, %) | 89 (36.2) | 45 (34.6) | 30 (34.9) | 14 (46.7) | 0.443 |
| Diabetes (N, %) | 42 (17.1) | 15 (11.5) | 19 (22.4) | 8 (26.7) | 0.040 |
| Barthel Index (median [25^th^–75^th^ percentile]) | 100 [100, 100] | 100 [100, 100] | 100 [100, 100] | 100 [100, 100] | 0.636 |
| Geriatric Depression Scale (median [25^th^–75^th^ percentile])  Depression (N, %) | 1 [0, 2]  10 (4.7) | 1 [0, 2]  3 (2.5) | 1 [0, 3]  6 (8.7) | 1 [0, 3]  1 (4.0) | 0.027  0.157 |
| Alcohol Use Disorders Identification Test (median [25^th^–75^th^ percentile])  Alcohol misuse (N, %) | 3 [1, 4]  14 (6.1) | 3 [1, 4]  5 (4.0) | 2 [1, 4]  5 (6.6) | 3 [1, 5.50]  4 (14.8) | 0.279  0.100 |
| ASA physical status (N, %)  1  2  3 | 19 (7.7)  151 (61.4)  76 (30.9) | 15 (11.5)  85 (65.4)  30 (23.1) | 3 (3.5)  50 (58.1)  33 (38.4) | 1 (3.3)  16 (53.3)  13 (43.3) | 0.020 |
| *Surgery characteristics* | | | | | |
| Surgical specialty (N, %)  Cardiothoracic  Intra-abdominal  Orthopedic  Other | 34 (13.9)  88 (36.1)  59 (24.2)  63 (25.8) | 18 (13.8)  50 (38.5)  29 (22.3)  33 (25.4) | 11 (13.1)  30 (35.7)  22 (26.2)  21 (25.0) | 5 (16.7)  8 (26.7)  8 (26.7)  9 (30.0) | 0.938 |
| Surgery duration, in minutes (median [25^th^–75^th^ percentile]) | 153 [94, 247] | 139 [90, 241] | 162 [95, 249] | 160 [111, 196] | 0.488 |
| Length of hospital stay, in days (median [25^th^–75^th^ percentile]) | 5 [3, 8] | 4 [2, 7] | 7 [3, 10] | 6.50 [4, 9] | <0.001 |
| Delirium (N, %) | 38 (15.4) | 16 (12.3) | 19 (22.1) | 3 (10.0) | 0.102 |
| Hospital mortality (N, %) | 2 (0.8) | 0 (0.0) | 2 (2.3) | 0 (0.0) | 0.153 |
| *3 months follow-up characteristics* | | | | | |
| Mortality before follow-up (N, %) | 5 (2.0) | 0 (0.0) | 5 (5.8) | 0 (0.0) | 0.009 |
| Trail Making Test A, in seconds (median [25^th^–75^th^ percentile]) | 42 [35, 52] | 41 [33, 50] | 42 [37, 52] | 46 [40, 53] | 0.170 |
| Trail Making Test B, in seconds (median [25^th^–75^th^ percentile]) | 85 [70, 112] | 82 [68, 106] | 98 [75, 123] | 88 [72, 112] | 0.076 |

*p-values of comparisons between the baseline + follow-up fMRI, the baseline fMRI only and the follow-up fMRI only group. Abbreviations: fMRI = funtional magnetic resonance imaging.

Supplemental Digital Content 6. Characteristics of the non-hospitalized controls and the total included sample.

|  | Non-hospitalized controls (N=50) | No delirium (N=208) | Delirium (N=38) | p |
| --- | --- | --- | --- | --- |
| *Baseline characteristics* | | | | |
| Center Utrecht (N, %) | 44 (88.0) | 107 (51.4) | 19 (50.0) | <0.001 |
| Female (N, %) | 20 (40.0) | 68 (32.7) | 17 (44.7) | 0.276 |
| Age (median [25^th^–75^th^ percentile]) | 71 [67, 75] | 70 [68, 74] | 73 [69, 75] | 0.111 |
| Mini Mental State Examination (median [25^th^–75^th^ percentile]) | 29 [28, 30] | 29 [28, 30] | 28 [27, 30] | 0.154 |
| Trail Making Test A, in seconds (median [25^th^–75^th^ percentile]) | 45 [37, 56] | 43 [36, 53] | 51 [40, 63] | 0.132 |
| Trail Making Test B, in seconds (median [25^th^–75^th^ percentile]) | 93 [81, 118] | 89 [72, 117] | 104 [74, 127] | 0.318 |
| Hypertension (N, %) | 17 (34.7) | 114 (55.6) | 20 (54.1) | 0.030 |
| Transient ischemic attack or stroke (N, %) | 5 (10.0) | 74 (35.6) | 15 (39.5) | 0.001 |
| Diabetes (N, %) | 10 (20.0) | 32 (15.4) | 10 (27.0) | 0.205 |
| Barthel Index (median [25^th^–75^th^ percentile]) | 100 [100, 100] | 100 [100, 100] | 100 [100, 100] | 0.450 |
| Geriatric Depression Scale (median [25^th^–75^th^ percentile])  Depression (N, %) | 1 [0, 2]  2 (4.0) | 1 [0, 2]  8 (4.5) | 1 [0, 2]  2 (5.9) | 0.608  0.917 |
| Alcohol Use Disorders Identification Test (median [25^th^–75^th^ percentile])  Alcohol misuse (N, %) | 3.50 [2, 4]  1 (2.0) | 3 [1, 4]  10 (5.1) | 2 [1, 4]  4 (11.8) | 0.137  0.144 |
| ASA physical status (N, %)  1  2  3 | 14 (28.0)  27 (54.0)  9 (18.0) | 17 (8.2)  128 (61.5)  63 (30.3) | 2 (5.3)  23 (60.5)  13 (34.2) | 0.001 |
| *3 months follow-up characteristics* | | | | |
| Mortality before follow-up (N, %) | 0 (NaN) | 5 (2.4) | 0 (0.0) | NaN |
| Trail Making Test A, in seconds (median [25^th^–75^th^ percentile]) | 37 [34, 49] | 42 [33, 51] | 42 [38, 62] | 0.311 |
| Trail Making Test B, in seconds (median [25^th^–75^th^ percentile]) | 84 [69, 106] | 83 [69, 110] | 95 [81, 117] | 0.277 |

*p-values of comparisons between the control, the no delirium and the delirium group.

Supplemental Digital Content 7. Consistency of fMRI measurements over time

To test consistency of the fMRI outcomes, changes in global functional connectivity strength, MST diameter and MST leaf fraction were analyzed within a non-surgical control group. The non-surgical control group was recruited via general practitioners in Utrecht and in Berlin (for demographic see Supplemental Digital Content 4). Inclusion criteria of non-surgical control participants were similar as the surgical group, with the exception of scheduled major surgery in the coming three months. A mixed model was constructed comparing the baseline and the follow-up measurement.

Consistency analysis in the non-hospitalized control group revealed no differences between baseline and follow-up of global functional connectivity strength (mean ±SD baseline = 0.77 ±0.03, mean ±SD follow-up = 0.77 ±0.03, β = 0.007, p = 0.175), MST diameter (mean ±SD baseline = 0.13 ±0.02, mean ±SD follow-up = 0.13 ±0.02, β = -0.002, p = 0.631), MST leaf fraction (mean ±SD baseline = 0.41 ±0.02, mean ±SD follow-up = 0.42 ±0.02, β = 0.003, p = 0.469), PCC-DLPFC-FC (left: mean ±SD baseline = 0.24 ±0.10, mean ±SD follow-up = 0.28 ±0.13, β = 0.035, p = 0.162; right: mean ±SD baseline = 0.25 ±0.11, mean ±SD follow-up = 0.26 ±0.13, β = 0.010, p = 0.719) over the two measurements.

Supplemental Digital Content 8. Mean values of the functional brain network outcomes at baseline and follow up for the delirium and the no delirium group.

| Functional brain network outcome | No delirium | Delirium |
| --- | --- | --- |
| Global functional connectivity strength baseline (mean ±SD)* | 0.76 ±0.03 | 0.77 ±0.03 |
| Global functional connectivity strength follow up (mean ±SD)* | 0.77 ±0.03 | 0.77 ±0.03 |
| MST diameter baseline (mean ±SD) | 0.13 ±0.02 | 0.14 ±0.03 |
| MST diameter follow up (mean ±SD) | 0.13 ±0.02 | 0.14 ±0.02 |
| MST leaf fraction baseline (mean ±SD) | 0.42 ±0.02 | 0.41 ±0.02 |
| MST leaf fraction follow up (mean ±SD) | 0.42 ±0.02 | 0.41 ±0.02 |
| Functional connectivity PCC – left DLPFC baseline (mean ±SD) | 0.25 ±0.10 | 0.23 ±0.11 |
| Functional connectivity PCC – left DLPFC follow up (mean ±SD) | 0.27 ±0.12 | 0.25 ±0.11 |
| Functional connectivity PCC – right DLPFC baseline (mean ±SD) | 0.26 ±0.11 | 0.25 ±0.12 |
| Functional connectivity PCC – right DLPFC follow up (mean ±SD) | 0.25 ±0.11 | 0.26 ±0.15 |

*Global functional connectivity strength was calculated by averaging the connectivity values of all connections in the minimum spanning tree (backbone network using the highest connectivity values), abbreviations: SD: standard deviation; MST: minimum spanning tree; PCC: posterior cingulate cortex; DLPFC: Dorsolateral prefrontal cortex.

Supplemental Digital Content 9. Associations between each of the functional network outcomes and delirium over the two fMRI measurements (i.e. preoperative and three months after surgery) obtained by mixed model analyses.

|  | *Global functional connectivity strength* | | | *Functional network efficiency (MST diameter)* | | | *Functional network integration (MST leaf fraction)* | | |
| --- | --- | --- | --- | --- | --- | --- | --- | --- | --- |
|  | β | 95% CI | p | β | 95% CI | p | β | 95% CI | p |
| (Intercept) | 0.664 | (0.577-0.751) | 0.000 | 0.113 | (0.047-0.178) | 0.001 | 0.421 | (0.351-0.492) | 0.000 |
| Timing of measurement | 0.006 | (0.001-0.010) | 0.021 | 0.000 | (-0.004-0.005) | 0.871 | -0.004 | (-0.008-0.001) | 0.135 |
| Delirium | 0.007 | (-0.003-0.016) | 0.165 | 0.004 | (-0.004-0.012) | 0.339 | -0.001 | (-0.009-0.008) | 0.844 |
| Age | 0.001 | (0.000-0.001) | 0.008 | 0.000 | (0.000-0.001) | 0.849 | 0.000 | (-0.001-0.000) | 0.200 |
| Gender | -0.009 | (-0.015--0.003) | 0.004 | 0.000 | (-0.005-0.004) | 0.895 | 0.005 | (0.000-0.010) | 0.035 |
| MMSE | 0.001 | (-0.001-0.003) | 0.424 | 0.001 | (-0.001-0.002) | 0.481 | 0.001 | (-0.001-0.003) | 0.444 |
| *Surgical specialty* |  |  |  |  |  |  |  |  |  |
| Cardiac surgery | ref | ref | ref | ref | ref | ref | ref | ref | ref |
| Intra-abdominal | 0.015 | (0.005-0.025) | 0.003 | 0.000 | (-0.008-0.007) | 0.911 | 0.000 | (-0.008-0.008) | 0.958 |
| Orthopedic | 0.004 | (-0.007-0.015) | 0.500 | -0.002 | (-0.011-0.006) | 0.563 | 0.001 | (-0.008-0.010) | 0.854 |
| Other specialty | 0.012 | (0.000-0.023) | 0.042 | -0.005 | (-0.014-0.003) | 0.232 | 0.002 | (-0.007-0.011) | 0.712 |
| Surgical duration in minutes | 0.000 | (0.000-0.000) | 0.907 | 0.000 | (0.000-0.000) | 0.273 | 0.000 | (0.000-0.000) | 0.917 |
| Center (Utrecht) | 0.011 | (0.005-0.017) | 0.001 | 0.007 | (0.002-0.012) | 0.004 | -0.010 | (-0.015--0.004) | 0.000 |
| Timing of measurement*Delirium | -0.015 | (-0.028--0.002) | 0.026 | 0.000 | (-0.013-0.013) | 0.977 | 0.002 | (-0.011-0.015) | 0.771 |

See next page for the associations of PCC-DLPFC-FC left and PCC-DLPFC-FC right.

|  | *PCC-DLPFC-FC left* | | | *PCC-DLPFC-FC right* | | |
| --- | --- | --- | --- | --- | --- | --- |
|  | β | 95% CI | p-value | β | 95% CI | p-value |
| (Intercept) | -0.026 | (-0.368-0.316) | 0.881 | 0.328 | (-0.046-0.702) | 0.085 |
| Timing of measurement | 0.022 | (-0.003-0.047) | 0.085 | -0.008 | (-0.033-0.017) | 0.520 |
| Delirium | -0.006 | (-0.050-0.037) | 0.777 | -0.005 | (-0.050-0.041) | 0.837 |
| Age | 0.001 | (-0.001-0.004) | 0.228 | 0.000 | (-0.003-0.002) | 0.764 |
| Gender | -0.004 | (-0.028-0.021) | 0.778 | -0.011 | (-0.038-0.015) | 0.402 |
| MMSE | 0.006 | (-0.004-0.015) | 0.244 | -0.002 | (-0.012-0.009) | 0.740 |
| *Surgical specialty* |  |  |  |  |  |  |
| Cardiac surgery | ref | ref | ref | ref | ref | ref |
| Intra-abdominal | 0.023 | (-0.015-0.062) | 0.231 | 0.040 | (-0.002-0.082) | 0.062 |
| Orthopedic | 0.010 | (-0.033-0.053) | 0.656 | 0.022 | (-0.025-0.069) | 0.365 |
| Other specialty | 0.009 | (-0.036-0.053) | 0.698 | 0.020 | (-0.029-0.068) | 0.420 |
| Surgical duration in minutes | 0.000 | (0.000-0.000) | 0.882 | 0.000 | (0.000-0.000) | 0.475 |
| Center (Utrecht) | -0.017 | (-0.043-0.009) | 0.190 | -0.001 | (-0.029-0.027) | 0.966 |
| Timing of measurement*Delirium | -0.003 | (-0.070-0.064) | 0.929 | 0.031 | (-0.036-0.098) | 0.365 |

* The β with 95% CI of timing of measurement reveal the change in functional network outcomes between the two measurements (preoperative and three months after surgery). For delirium, the β with 95% CI indicates differences in functional network outcomes at the preoperative measurements. The interaction term between these two presents whether change in functional network outcomes between the two measurements was different between delirious and non-delirious patients. A random intercept was used for each individual. β: beta coefficient, MST: minimum spanning tree, CI: confidence interval, ref: reference.
